# Adverse effects of antipsychotic drugs on metabolism depend on drug dosing and feeding times

**DOI:** 10.1101/2022.02.23.22271365

**Authors:** Rizaldy C Zapata, Allison Silver, Dongmin Yoon, Besma Chaudry, Avraham Libster, Michael J McCarthy, Olivia Osborn

## Abstract

Antipsychotic drugs (AP) are highly efficacious treatments for psychiatric disorders but are associated with significant metabolic side effects. The circadian clock maintains metabolic homeostasis by sustaining daily rhythms in feeding, fasting and hormone regulation but how circadian rhythms interact with AP and its associated metabolic side effects is not well known. In these studies, we investigated the impact of time of AP dosing on the development of metabolic side effects. In mice, AP dosing at the start of the light cycle (AM) resulted in significant increase in food intake, weight gain compared with equivalent dose before the onset of darkness (PM). Time of AP dosing also impacted circadian gene expression, metabolic hormones and inflammatory pathways and their diurnal expression patterns. To examine the possibility of time-dependent AP effects in humans, we conducted a retrospective examination of weight and metabolic outcomes in patients who received risperidone (RIS) for the treatment of serious mental illness. Using pharmacy records to estimate the time of RIS dosing, we observed a significant association between time of dosing and severity of RIS-induced metabolic side effects. Eating within a restricted time window (Time restricted feeding/eating, TRF/TRE) has been shown in both mouse and human studies to be an effective therapeutic intervention against obesity and metabolic disease. We demonstrate, for the first time, that TRF is an effective intervention to reduce AP-induced metabolic side effects in mice. These studies identify highly effective and translatable interventions to mitigate AP-induced metabolic side effects.

## INTRODUCTION

Antipsychotic drugs (AP) are highly efficacious treatments for serious mental illnesses (SMI) such as schizophrenia, bipolar disorder and major depression, but frequently cause weight gain and metabolic disease (1). The metabolic side effects of APs exacerbate medical comorbidities and worsen health outcomes in SMI patients, a population already at high risk for cardiometabolic disorders and early mortality(2). Moreover, AP-induced metabolic side effects lead to treatment discontinuation and non-compliance that destabilize SMI and worsen outcomes such as social/occupational impairment and suicide. Both neurobehavioral and peripheral metabolic factors have been implicated as key factors driving adverse effects of AP but the specific molecular mechanisms remain unknown (3). Previous studies have identified potential adjuvant drugs that can mitigate some of the weight gain effects of APs (4-10) but these interventions have yet to make a significant impact on the problem associated with APs. Additional strategies are needed to further reduce the metabolic burden of AP in patients with SMI to make these drugs safer, more tolerable but still effective.

Circadian rhythms are controlled by multiple centers including the central clock in the hypothalamic suprachiasmatic nucleus (SCN) that regulates food intake and behavior while the peripheral clocks in the liver, adipose, pancreas, muscle and other metabolically active tissues control energy expenditure and whole-body insulin sensitivity (11). Central and peripheral clocks in cells generate circadian rhythms to optimize energy harvest and utilization across the 24-hour dark/light cycle. A transcriptional/ translational feedback loop made up of ∼20 “core clock genes” maintains essential functions underlying cellular circadian rhythms. At the center of this loop, the proteins Clock and Bmal1 bind to form a heterodimeric transcriptional activator. The Bmal1 complex drives the expression of period (*Per1/2*) genes, and cryptochrome (*Cry1/2*) transcriptional repressors that sustain circadian oscillators with a period length of ∼24-hrs. A second feedback loop provides additional robustness to the oscillatory mechanism and consists of nuclear receptors of the Rev-erb*α* which drives rhythmic *Bmal1* expression. These intricate feedback loops generate rhythms with a period of about a day that maintains metabolic homeostasis by sustaining daily rhythms in feeding and fasting(12)ating within a restricted time window (Time Restricted Feeding (TRF) has been shown in both mouse and human studies to be an effective therapeutic intervention against obesity and metabolic disease (12, 13). However, to date there has been very little consideration of how circadian rhythms, timing of feeding and APs may interact to affect metabolism and behaviors governing weight.

In our previous work, we found that the timing of administration of the AP sulpiride affected the weight gain and metabolic outcomes in mice (14). Sulpiride acts by selective antagonism of the D2 dopamine receptor whereas most commonly used AP have a more complex mechanism involving additional receptors and are associated with greater risk for weight gain. While almost all APs result in weight gain (15), the “second generation” APs such as risperidone (RISP) and olanzapine (OLZ) cause the most weight gain (16, 17) and are widely prescribed (18). In these studies, we evaluated whether dosing time of OLZ or RISP impacted AP-induced weight gain and metabolic side effects. We then further explored whether TRF can mitigate the metabolic impairments associated with AP-induced weight gain.

## RESULTS

### Time of dosing impacts AP-induced metabolic side effects in mice

To investigate the impact of time of AP dosing on metabolic side effects, we used a mouse model of AP-induced hypeprhagia and weight gain. Dosing of APs at a specific time of day was achieved through self-administration of AP (RIS or OLZ) mixed into peanut butter pellets (19). AP treatment at the start of the light cycle (AM) resulted in significantly higher food intake (**Fig. 1A,B**) and weight gain (**Fig. 1C,D**) compared with CON treatment. In contrast, APs administed at the start of the dark cycle (PM) did not induce significant increases in food intake and body weight compared with CON treatment at that time. Therefore, the hyperphgaia and weight gain side effects of APs are highly depedent on the time of dosing. Two hours after AP treatment, we measured circulating blood glucose and observed increased glucose in RIS-AM (**Fig. 1E**) treated mice compared with RIS-PM or placebo-control groups. No significant changes in glucose were observed with OLZ (**Fig. 1F**). RIS is the more commonly prescribed than OLZ and thus we focused on this group in the detailed follow up studies. RIS-AM dosing also resulted in significantly higher liver weight (**Fig. 1G**) and increased levels of plasma triglycerides (**Fig. 1H**) comapred with RIS-PM further suggesting that timing of dosing across the light/dark cycle significantly impacts metabolic side effects induced by APs.

**Figure 1.**
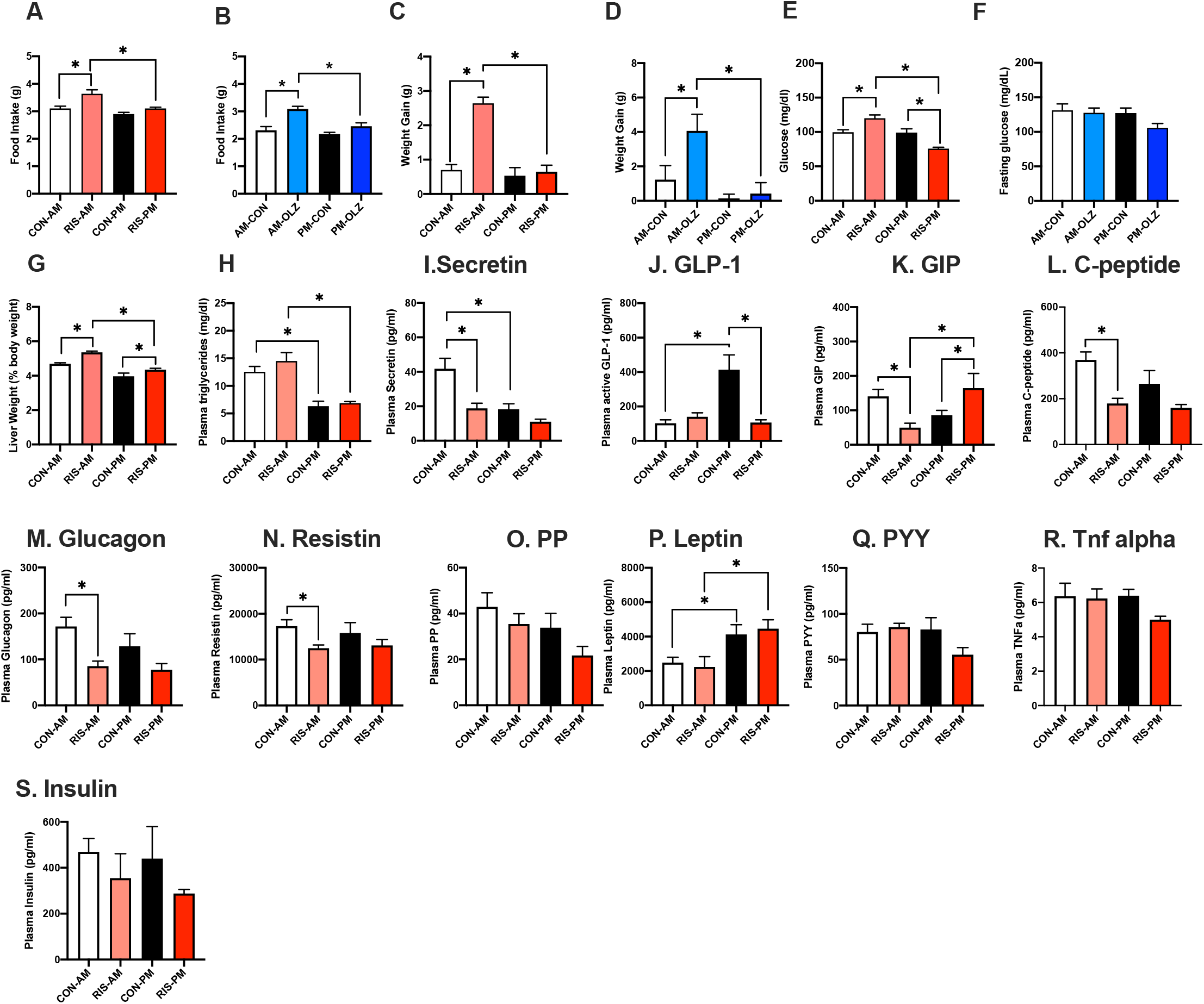
Timing of AP dosing impacts metabolic side effects in mice. A-B Food intake, C-D. weight gain and (E-F) fasting glucose (2 hours post-treatment) of mice treated with risperidone (RIS, 3mg/kg, A,C,E) or olanzapine, (OLZ, (8 mg/kg, B,D,F) during the light (ZT2) or dark cycle (ZT11). G. liver weight, Plasma H. Triglyceride levels. I. Secretin, J. Glucagon-Like Peptide 1, K. Gastric Inhibitory Peptide, L. C-peptide, M. Glucagon, N. Resitin, O. Pancreatic Polypeptide, P. Leptin, Q. Peptide YY, R. TNF-α, and S. Insulin levels in mice treated with risperidone (3 mg/kg) during the light (ZT2) or dark cycle (ZT11). Data is expressed as mean ± SEM and analyzed using 2-Way ANOVA followed by Fisher’s LSD posthoc test. Significance was set at *p* < 0.05.

Many metabolic hormones are rhythmically released and regulated by the circadian clock. For this reason, we evaluated whether the changes in food intake and metabolism are associated with dysregulated hormone secretion. The significant effects of RIS on secretin **(Fig. 1I)**, GLP-1 **(Fig. 1J)** and GIP **(Fig. 1K)** are highly depedent on the time of dosing. RIS-AM dramatically decreased secretin to similar levels seen in CON-PM despite secretin levels being twice as much in CON-AM. RIS-AM treatment abolished the reduced GLP-1 levels seen between RIS-PM and CON-PM when compared to CON-AM. Interestingly, there is a potential phase shift with Gastric Inhibitory Polypeptide (GIP) with RIS-AM decreasing GIP to level comparable to CON-PM while RIS-PM increased GIP to CON-AM levels. Although RIS treatment in general decreased C-peptide **(Fig. 1L)**, Glucagon **(Fig. 1M)** and Resistin **(Fig. 1N)**, the effect of RIS-AM is more evident than RIS-PM when compared to their respective placebo controls. Pancreatic Polypeptide (PP) **(Fig. 1O)** was decreased, and Leptin **(Fig. 1P)** was increased during PM with no significant RIS effect. No significant effects of drug and time of dosing were observed with Peptide YY (PYY) **(Fig. 1Q)**, TNF-α **(Fig. 1R)** and Insulin **(Fig 1S)**. Therefore, overall, timing of AP dosing had significant effects on some metabolic hormone levels.

### Time of dosing of APs significantly effects circadian gene expression

To examine the effects of APs on the expression of circadian genes process, we evaluated circadian gene expression at two time points (AM/PM) in the liver, hypothalamus and gonadal white adipose tissue (gWAT) of CON and RIS-treated mice. In the hypothalamus **(Fig. 2A)**, consistent with the known patterns of variation across time, there is significant temporal differences on the gene expressions of many of these genes. For example, *Per1, Per2* and *Cry1* expressions were increased during the PM with *Rev-erbα* and *Bmal1* expressions were lower at this time compared with CON-AM. The effects of RIS on hypothalamic *Clock* and *Cry2* expression were time dependent. RIS-AM increased *Clock* expression while RIS-PM increased *Cry2* expression compared to their respective controls. Peripheral tissues such as liver **(Fig. 2B)** and gWAT **(Fig. 2C)** demonstrated similar temporal profiles of circadian gene expression. In both tissues, the expression of *Per1, Per2, Cry1* and *Cry2* increased during the PM while *Rev-erbα* expression was lower at this time compared with CON-AM. Both tissues also responded similarly to time-dependent effect of RIS - with RIS-AM decreasing *Rev-erba* compared to CON-AM with no changes seen between CON-PM and RIS-PM. However, there are some tissue differences observed. For example, comparison of *Clock* expression in the CON groups revealed oppostite trends in liver and gWAT, wherby *Clock* expression in the CON-PM group is lower in the liver, but higher in the gWAT compared with the CON-AM group. RIS-AM treatment decreased *Clock* expression in the liver while RIS-PM increased it in the gWAT compared with CON-AM groups. Furthermore, RIS-AM significantly lowered liver expression levels of *Clock, Per1, Per2, Cry2*, and *Rev-erbb* compared with CON-AM but these genes were unchanged in the gWAT in RIS-AM versus CON-AM groups. In contrast, *Clock, Per1* and *Cry2* expression were differentially expressed with RIS-PM compared wih CON-PM in the gWAT but were unchanged in the liver. In addition, gWAT *Bmal1* expression was lower in RIS-PM compared to RIS-AM but unchanged in the liver. Overall, transcriptional analysis of the liver, gWAT and hypothalamus revealed RIS treatment induces significant changes in circadian gene expression in multiple tissues that is further perturbed by dosing in the AM. While the temporal resolution of these studies is limited, these time- and tissue depedent differences in circadian gene expression indicate that there may be widepread change in the circadian rhythm phase and/or amplitude caused by RIS in the gWAT and liver.

**Figure 2.**
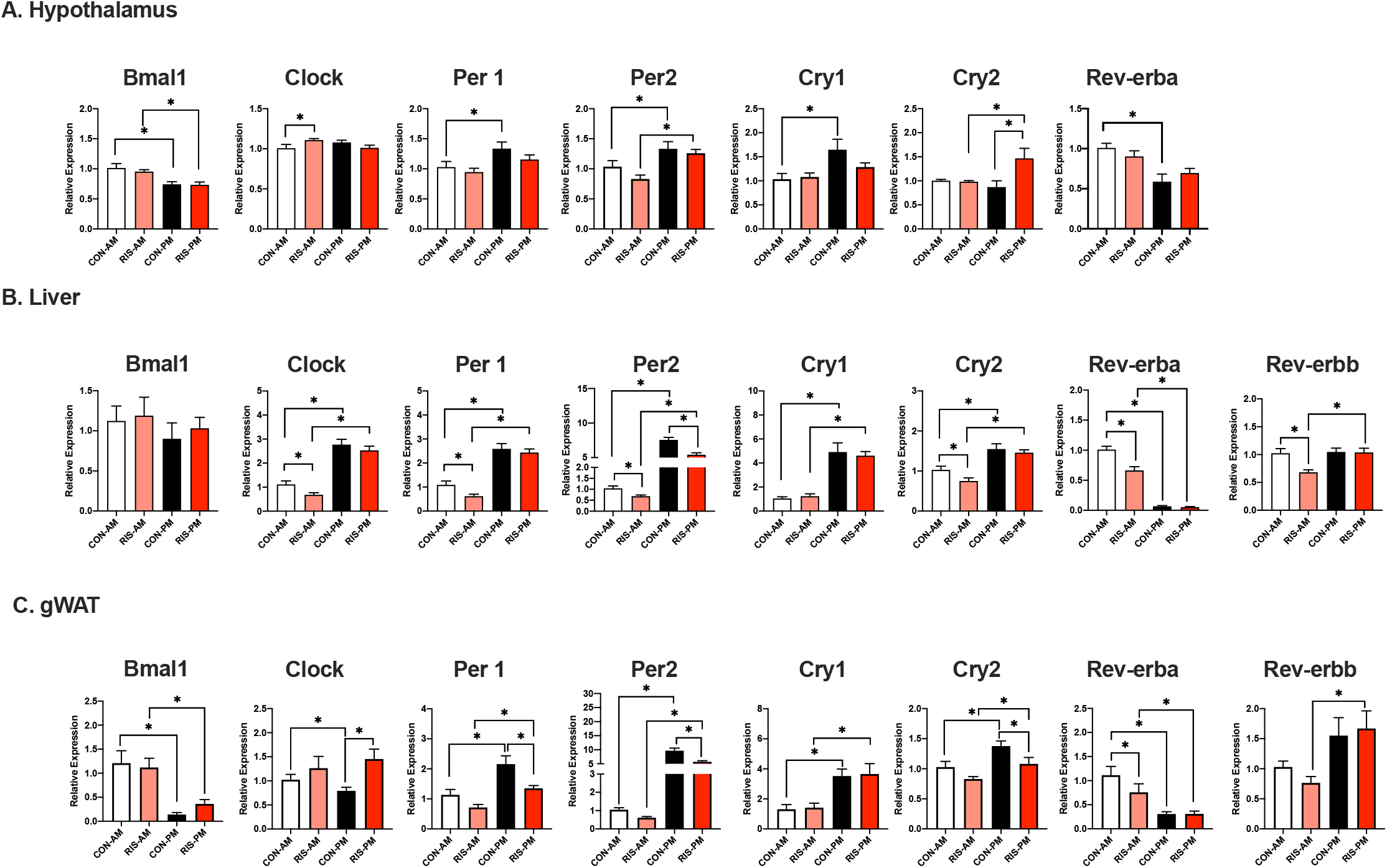
Effect of the timing of RIS dosing on circadian gene expression in the metabolic tissues. Circadian Gene Expression in A. Hypothalamus, B liver, C. gWAT of mice treated with risperidone (RIS, 3 mg/kg) or placebo control (CON) during the light (AM = ZT2) or dark cycle (PM =ZT11). Data is expressed as mean ± SEM and analyzed using 2-Way ANOVA followed by Fisher’s LSD posthoc test. Significance was set at *p* < 0.05.

### Time of dosing of APs significantly effects inflammatory gene expression

Our recent study identified that inflammation potentiates AP-induced hyperphagia and weight gain (20). Since inflammation is regulated in part by the circadian clock (21), we evaluated whether the timing of AP dosing affects inflammation in metabolically active tissues (hypothalamus, liver and gWAT). Hypothalamic expression of inflammatory markers did not change between CON-AM and CON-PM groups, but RIS AM dosing was assocaied with higher levels of proinflammatory genes including *IL-1ß, TNFα* and *Cd11c* compared with RIS-PM dosing. In the liver, the majority of genes were also unchanged by CON-AM vs CON-PM dosing, with the exception of *TNF-α* that was higher in CON-PM compared with CON-AM. *TNF-α* was also differentially expressed between RIS-AM and RIS-PM groups (**Fig. 3B**). *Cd11b*, a marker of myeloid cells was elevated in the liver by both RIS-AM/PM treatment groups compared with respective CON groups. Liver *IL-1ß* levels followed a similar pattern as observed in the hypothalamus whereby IL-1ß levels were elevated in both AP treatment groups compared with respective CON groups (**Fig. 3B**). Expression levels of *Cd11c*, a marker of proinflammatory macrophages, were elevated in the liver of RIS-AM treated mice compared with CON-AM but not different betweeen RIS-AM and RIS-PM groups. Levels of *IL-10*, an anti-inflammatory cytokine were significantly lower in RIS-AM treated mice compared with RIS-PM. Inflammatory gene expression in gWAT was unchanged between CON-AM and CON-PM groups. RIS-AM treatment resulted in higher inflammatory tone in the gWAT compared with RIS-PM dosing. In particular, *Tnf-α* and *Cd11c* and *IL-10* levels were increased in RIS-AM compared to RIS-PM (**Fig. 3C**). Therfore, RIS-AM dosing resulted in signifcant changes in inflamatory tone across multiple metabolic tissues including liver, gWAT and the hypothalamus.

**Figure 3.**
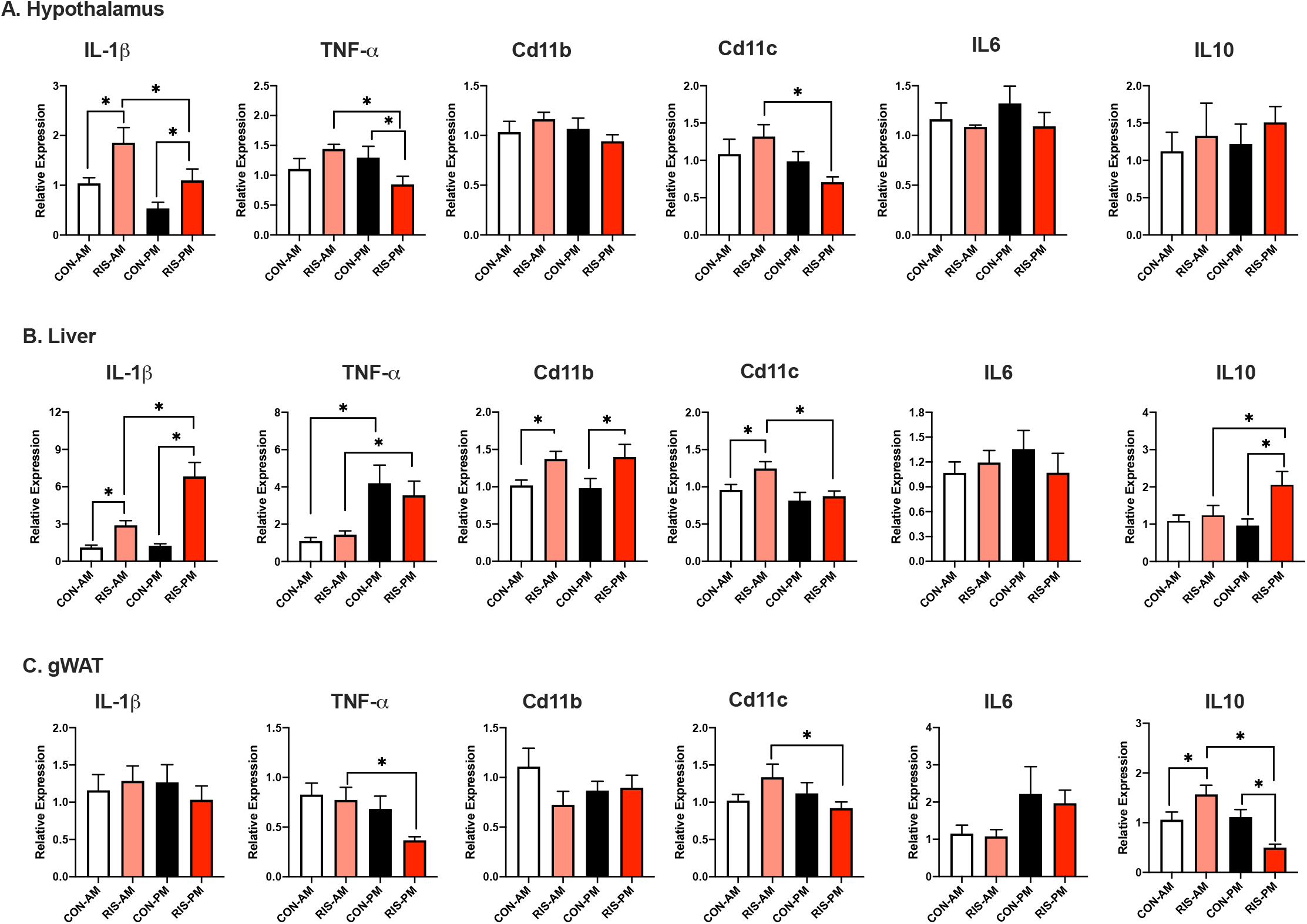
Effect of the timing of RIS dosing on inflammatory gene expression in the metabolic tissues. Inflammatory Gene Expression in A. Hypothalamus, B liver, C. gWAT of mice treated with risperidone (RIS, 3 mg/kg) or placebo control (CON) during the light (AM = ZT2) or dark cycle (PM =ZT11). Data is expressed as mean ± SEM and analyzed using 2-Way ANOVA followed by Fisher’s LSD posthoc test. Significance was set at *p* < 0.05.

### Time of dosing impacts AP-induced metabolic side effects in humans

To examine the possibility of time-dependent AP effects in humans, we conducted a retrospective examination of weight and metabolic outcomes in patients who received RIS for the treatment of SMI using pharmacy records to estimate the time of RIS dosing. After controlling for treatment compliance, demographic and clinical factors, patients taking RIS-PM before the onset of sleep for ∼1 year gained significantly more weight (**Fig. 5A**), had elevated glycosylated hemoglobin A1c (HbA1c) (**Fig. 5B**) compared to patients taking RIS-AM upon waking in the morning. These results are opposite to the differences observed in mice in terms of AM/PM, but similar to mice in terms of the relationship between AP dosing to sleep/activity schedules. No changes were determined in total plasma cholesterol, LDL, HDL and triglycerides (**Fig. 5C-F**).

### TRF modulated the weight gain side-effect of RIS-AM treatment

Time restricted feeding (TRF) without calorie restriction is a therapeutic intervention against obesity and insulin resistance in both mouse and human studies(22, 23). Therefore, we tested whether TRF could mitigate RIS-induced metabolic impairments. Since the RIS-induced weight gain and impairments were most pronounced in the RIS-AM group, we used this dosing scheduling in TRF studies. As expected, RIS treatment in *ad libiutm* fed mice increased food intake (**Fig. 5A**), weight gain (**Fig. 5B**) and blood glucose levels (**Fig. 5C**) compared with CON treated *ad libitum* fed mice, Importantly, TRF significantly mitigated RIS-induced food intake (**Fig. 5A**), weight gain (F**ig. 5B**) and lowered blood glucose (**Fig. 5C**) compared with the *ad libitum* fed RIS treated group. Evaluation of expression of the core circadian genes revelaved key RIS-induced expression changes in Liver *Per1* (**Fig. 4D**), gWAT *Clock* (**Fig. 4E**) and *Cry1* (**Fig. 4F**) were ‘rescued’ by TRF to equivalent CON expression levels. Furthermore, AP-induced inflammatory gene expression changes were also attenuated by TRF treatment (**Fig 4G-K**). For example, *Il-1ß* levels were consistently elevated by RIS treatment compared with CON treatment in *ad libitium* fed mice, and TRF speficially reduced RIS-induced *IL-1ß* levels in hypothalamus (**Fig. 4G**) liver (**Fig 4H**) and adipose tissue (**Fig. 4I**) compared with RIS-treated mice on the *ad libitium* diet. Other inflammatory markers, such as *Il-6* (**Fig. 4J**) and *Tnf-α* (**Fig.4 K**) were also reversed by TRF in the hypothalamus.

**Figure 4.**
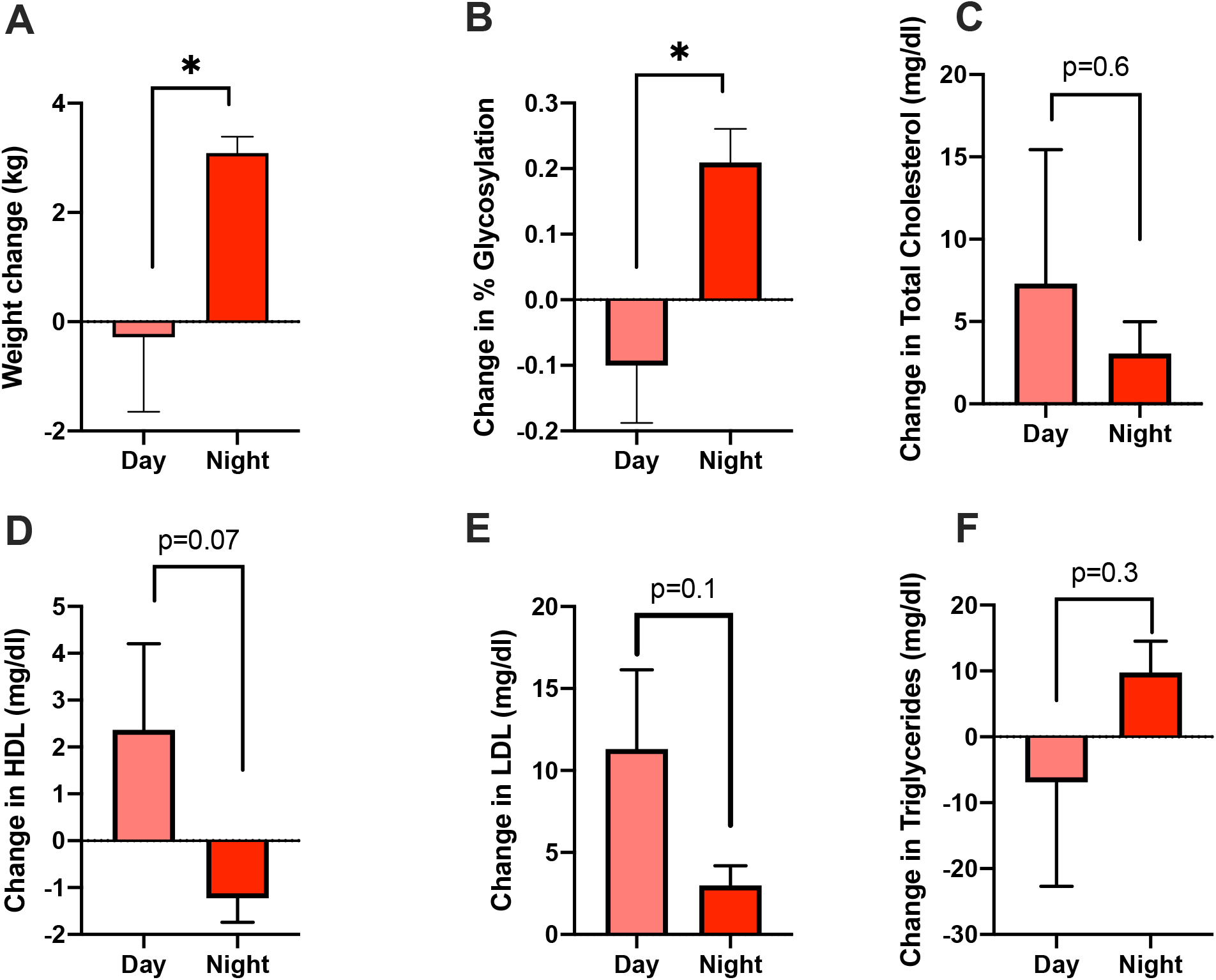
Taking RIS at night increased weight gain and worsen glycemic index compared with morning dosing. Changes in A. Body weight B. HbA1c, C. Total cholesterol, D. Change in low-density lipoprotein (HDL) E. Change in low-density lipoprotein (LDL), F. Change in plasma Triglycerides, of patients taking risperidone (RIS) either during the daytime or nighttime using pharmacy records from the VA San Diego Healthcare System. Data is expressed as mean ± SEM and analyzed using Student T-test. Significance was set at *p* < 0.05.

**Figure 5.**
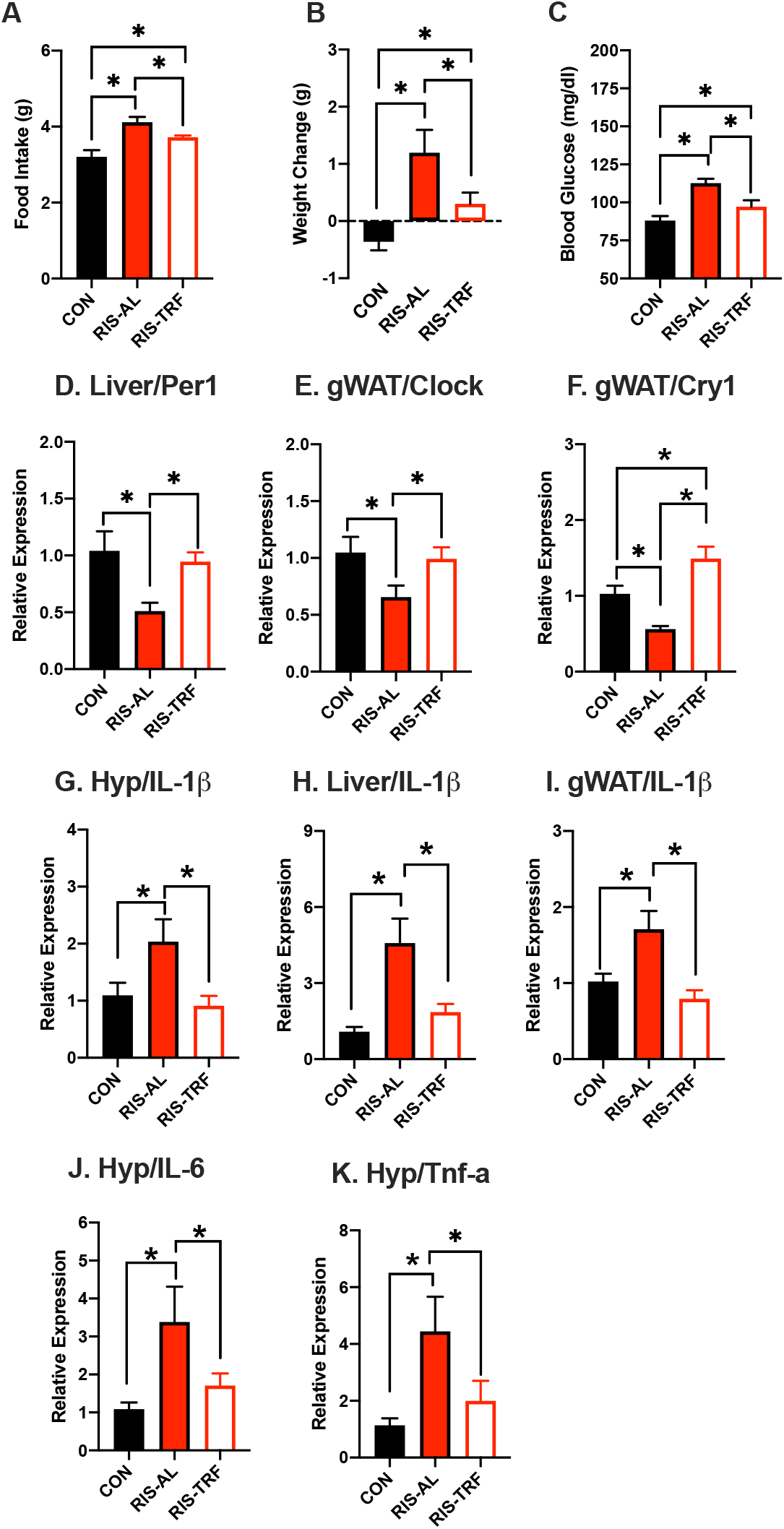
TRF mitigated the metabolic side-effects of RIS. A. Food intake, B. Body weight change, C. Blood glucose (2 hours post-treatment), (D-F) Circadian gene expression and (G-K) Inflammatoy gene expression in metabolic tissues, from mice treated with RIS during the light period with 24 hour food access compared with mice treated with RIS with only 12 hour food access. Data is expressed as mean ± SEM and analyzed using one-way ANOVA followed by Fisher’s LSD posthoc test. Significance was set at *p* < 0.05.

## DISCUSSION

These studies investigated the impact of time of AP dosing on the development of weight gain and metabolic side effects. In nocturnal mice, AP dosing at the start of the less active light period (AM) resulted in significant increase in food intake, weight gain and metabolic side effects compared with equivalent dose just before the start of the active dark period (PM). Time of AP dosing also impacts circadian gene expression, metabolic hormones and inflammatory pathways and their diurnal expression patterns. We also observed an association between time of dosing and severity of RIS-induced metabolic side effects in patients with SMI prescribed APs. We demonstrate, for the first time, that TRF is an effective intervention to reduce weight gain effects of APs in mice.

Circadian rhythms influence feeding behavior, activity, and body weight with significant impacts on metabolism. Our studies revealed that AP-treatment impacted circadian gene expression across multiple metabolic tissues. Liver expression of *Clock, Per1, Per2, Cry2* and *Rev-erb*α and gWAT expresison of *Per1, Per2, Cry2*, and *Rev-erbα* were lower in mice dosed with RIS in the AM compared with placebo control doisng at the same time. In support of the role of these genes impacting food intake, mice with whole body deletion of *Per1/2* or *Cry1/2* (24), or hypothalmic KO of Rev-erba/Rev-erbb (25) eat more during the light period and less during the dark period compared with WT mice. Therefore, RIS induced decreases in these circadian genes during the inactive light period may contribute to the AP-induced hyperphagia seen in this group. The hypothalamic SCN clock is often resilient to perturbaitons by environmental inputs (26) thus the impact of APs on hypothalamic circadian gene expression was more limited.

Circadian regulation of hormones including insulin, glucagon, leptin and cortisol (27, 28) facilitate metabolic coordination and adaptation to periods of feeding and fasting and is a feature of optimal physiological function (29). For example, insulin levels follow a regular circadian rhythm in humans, peaking during the day and dropping at night and disruption of molecular circadian rhythms causes insulin resistance and elevated blood glucose (11, 30). Prior studies have also shown that APs impact peripheral organs and dysregulate hormone regulation and metabolsim (31). Our data suggest RIS affects the physiological rhythms that coordinate metabolic hormones with feeding, fasting and activity behaviors in mice and humans. For example, RIS-AM treatment induced relative hyperglycemia compared with CON-AM or RIS-PM dosing. In addition, RIS-AM treatment also flattened the temporal profiles in blood levels of glucagon, GIP and secretin levels with losses of the diurnal peaks observed in CON mice. RIS-AM also significantly lowered C-peptide compared with placebo CON and a similar trend of lower insulin in RIS-AM vs CON-AM was observed, which may contirbute to the higher levels of glucose in RIS-AM vs CON-AM groups. Similar indications of sub-optimal metabolic function caused by RIS was shown by differences in inflammatory cytokines and changes in their diurnal profile, indicating metabolic stress response particularly in the liver and to some extent hypothalamus, especially in the RIS-AM group. Taken together, these results indicate that, in additon to quantitative differences in hormone levels caused by RIS-AM, the drug also strongly affects the temporal coordination among hormone regulators of feeding behavior, blood glucose and metabolic activity depending on when the drug is administered. Furthermore, loss of temporal coordination among these regulators in RIS treated animals is associated with systemic inflammation and with more weight gain. The timing of our sampling limits our ability to distinguish phase and amplitude effects or to detect subtle differences in timing. Repeated sampling at intervals throughout the day will be important to described the effects of RIS on the oscillation of these genes over a 24 h cycle.

Importantly, the rest-activity cycle is reversed between rodents (nocturnal) and humans (diurnal). Our studies in both species align in that APs taken at the start of the rest period (AM in rodents, PM in humans) exacerbates the metabolic side effects compared with dosing at the start of the active period. Retrospective examination of pharmacy records in SMI patients who received RIS revealed that patients taking the medication at night gained significantly more weight and had elevated HbA1c compared to patients taking RIS in the morning. Importantly, these findings extend our published work reporting similarly unfavorable lipid profiles in SMI patients taking aripiprazole at night (32). These data imply shifting the time of AP dose may be a clinical strategy to mitigate weight gain and metabolic side effects. However, on a practical level, APs also have a sedative effect that may pose challenges if taken at the start of the day. Thus, dosing at the start of the rest period while introducing time restricted eating could represent a highly effective alternative strategy that could be implemented to mitigate AP-induced side effects regardless of timing. When the consumption of high fat diet is restricted to an 8-9 hr window during the active phase, TRF animals are protected from the adverse metabolic consequences, even though the total number of calories consumed was similar (12, 22). In our study, TRF in the 12 hr active / dark phase abrogated RIS-induced weight gain and resulted in lower glucose levels compared with RIS treated *ad libitum* fed mice. TRF intervention in the RIS-AM group also resulted in 10% reduction in food intake which may have contributed to the reduced weight gain and overall improvements in glycemia. TRF was a highly effective intervention to reduce metabolic side effects in mice and it rescued AP-induced gene expression changes of key circadian and inflammatory genes in multiple metabolic tissues. For example, TRF restored liver expression of *Per1*, gWAT *Clock* and *Cry1* expression to equivalent levels observed in the *ad libitum* control groups. With only two time points, our findings cannot determine whether TRF caused quantitative changes in circadian gene expression and/or normalized the phase of rhythmic expression profiles of circadian genes. In the present context, either outcome could be regarded as potentially beneficial and are in line with a role for TRF restoring rhythms in models of circadian disruption(22). TRF also restored hypothalamic, liver and gWAT *Il-1ß* gene expression to equivalent levels observed in the *ad libitum* control groups. *IL1ß* is a proinflammatory cytokine with well described roles in metabolic disease (33) and neutralization of I*L1ß* improves glycemia and metabolic health in mouse studies (34). Whether *Il-1ß* is causative of RIS-induced metabolic side-effects or consequence of increased body weight warrants further studies. Recent studies have shown that TRF has metabolic benefits even in animals that lack a circadian rhythm (12). For example, when provided access to food ad libitum, whole-body *Cry1/Cry2* and in liver-specific *Bmal1* and *Rev-erbα/ß* knockout mice rapidly gained weight and showed genotype-specific metabolic defects. However, when fed the same diet under TRF (food access restricted to 10 hr during the dark phase), they were protected from excessive weight gain and metabolic diseases. This suggests that the TRF is sufficient to properly entrain metabolic pathways that would be otherwise dysregulated by the lack or perturbed circadian rhythm or in psychiatric populations where internal clock may be compromised (35, 36). Time-restricted eating also mitigates weight gain and metabolic disease in humans (23, 37-39).

In summary, we provide evidence that strongly suggests that APs dosed at the wrong time in the circadian cycle perturb the temporal coordination of circadian and metabolic regulators to cause significant effects on weight gain and metabolic health in both mouse and human. In humans, retrospective pharmacy records-based data demonstrate worse effects of APs at night on weight and glycemia. While compelling, future randomized, prospective clinical studies are needed to fully examine the impact of timing on AP-induced metabolic health. TRE represents an exciting strategy to mitigate AP-induced metabolic side effects. However, patient compliance with TRE in the SMI population may be difficult as these patients often suffer from irregular sleep and activity patterns independently of AP treatment (36). Moreover, APs can cause sleep disturbances (40) with some patients reporting increased frequency of night eating after taking RIS (41, 42), OLZ (43) and other APs (44, 45). In some patients, dosing Aps before the rest period may perturb the circadian rhythm to drive aberrant feeding behavior. Therefore, clinical studies are necessary to test if TRE is an effective intervention to mitigate AP-induced metabolic side effects in patients.

## MATERIALS AND METHODS

### Mouse studies

All procedures were approved by the University of California San Diego IACUC. Female C57BL6/J mice were purchased from The Jackson Laboratory (Stock number: 000664, Sacramento, CA) at 9-10 weeks of age. Mice were acclimatized to the laboratory conditions for 7 days. Animals were maintained in a 12-hour light:dark cycle with a humidity between 60–70%. At age 10-11 weeks mice were singly housed, provided unlimited access to water and normal chow food, and continued on a 12h/12h light dark schedule. Female mice were studied because they are particularly susceptible to AP-induced weight gain and reflect what is seen in both male and female SMI patients.

### Light and Dark AP Dosing

APs OLZ (8mg/kg) or RISP (3mg/kg) were self-administered to mice in a peanut butter/drug mixture pellet or placebo control peanut butter alone. Mice were trained to eat peanut butter by fasting overnight followed by introduction of a peanut butter pellet (placebo control). Placebo control pellets were then given to unfasted mice for an additional 3 consecutive days to overcome any novelty-associated behavior changes in feeding and locomotion. After training, mice consumed the pellet in less than ∼15 mins (20) allowing for precisely timed AP dosing. Dosing time was approximately 2 h after lights on between 8:00-8.30AM, (Zeitgeber time (ZT) 2) for the ‘AM’ group and between 5:30-6:00PM, ZT 11 for the ‘PM’ group. No changes were made to the light/dark cycle. At the end of the two-week study, mice were sacrificed 2 hrs after the last AP dose. AM groups were dosed at 8:00AM and sacrificed at 10:00AM while PM groups were dose at 6:00PM and were sacrificed at 8:00 PM, n=7-8 per group. Blood and tissues were dissected, flash frozen with liquid nitrogen and stored at -80°C until analyses.

### Time Restricted Feeding

Since the weight gain and metabolic effects of AP were only observed during the AM administration, we designed the TRF study using RIS-AM only. Mice were divided into 3 groups (CON-*ad lib*, RIS-AL, RIS-TRF, n=6-8 per group). We did not include a CON-TRF group as previous studies have investigated the impact of TRF on body weight and metabolism (12, 22). Water was freely available at all times. In the CON ad lib and RIS-AM-ad lib groups, food was available 24 h a day. In the TRF group, food access was removed between ZT0 and ZT12 and restored during the active phase (ZT12-24) using specifically designed cages with a rotating wheel designed to have slots loaded with food accessible only at scheduled times. Food intake was measured daily at ZT0 and ZT12 and body weight measured at ZT0. After 10 days, mice were sacrificed 2hr after AP dosing (dosed at 8AM, sacrificed at 10AM).

### RNA extraction, Quantitative PCR and Determination of metabolic hormone levels

Total RNA was extracted using Trizol (Invitrogen) and RNeasy Extraction Kit (Qiagen) as recommended by the manufacturer. RNA concentration and quality were assessed using Nanodrop. cDNA was synthesized from 500 ng of RNA using High-Capacity cDNA transcription kit (Thermo Fisher). qPCR was performed using StepOne Plus (Applied Biosciences). Gene expression was normalized to housekeeping genes acidic ribosomal phosphoprotein P0 (36B4) for the liver (46), ATP synthase F1 subunit epsilon (Atp5e) for both gonadal and brown adipose tissues (47) and hypoxanthine phosphoribosyltransferase 1 (Hprt1) for the hypothalamus (48). These genes are stable reference genes suitable for circadian studies in different tissues and mouse strains (46, 49). Our data also confirm no difference in the expression of these housekeeping genes between AM and PM placebo groups (Supplemental Figure 1) Circulating blood levels of cytokines and gut-derived hormones were measured by multiplex ELISA (MMHE-44k-15, Millipore Sigma, Burlington, MA, USA).

### Human retrospective analysis

Using pharmacy records from the VA San Diego Healthcare System (VASDHS), we conducted a retrospective study of timing, weight gain, and long-term metabolic outcomes in SMI patients (age 18-75 years) taking RIS for approximately 1 year. The start date was defined as the first day that RIS was released to the patient and the end date was defined as the start date plus 365 days. Measures of body weight, HbA1c, serum glucose, fasting lipids were included if the first measure preceded initiation of RIS and the interval between the first and second measures was 9-15 months (1 year ±3 months). Patients were considered to have taken RIS in the morning if the instructions indicated “morning”, “daily”, “qam” or “qday” and were considered to have taken RIS at night when the instructions indicated “qhs” or “bedtime”. Compliance of > 0.8 over 1 year was estimated by the frequency of on time refills. Data from January 1, 2002 to December 31, 2016 were included in the analysis. Those taking ultra-low dose RIS (≤0.25 mg), formulations of injectable RIS or RIS concurrent with another AP were excluded. Clinical indications for RIS included schizophrenia, schizoaffective disorder, unspecified psychosis, bipolar disorders, depressive disorders, and post-traumatic stress disorder. Patients with a diagnosis of Alzheimer’s disease, Parkinson’s disease, dementia or other neurocognitive disorder were excluded. Subjects with existing metabolic disorders prior to starting RISP including T2D, essential hypertension, and hypercholesterolemia were excluded. For each variable, data were analyzed using ANCOVA comparing the change in metabolic parameters from start date to end date in the AM vs. PM group, with age, sex, race, and dose of RISP as covariates. Smoking, alcohol and substance use history were not reliably recorded in the pharmacy database and were not considered. The research was reviewed and approved by the VASDHS IRB. Body weight data was available for n=102 “morning”, n= 2014 “evening” dosing, A1c data was available for n=42 “morning”, n= 303 “evening” dosing, LDL data was available for n=39 “morning”, n= 727 “evening” dosing, HDL, TG, Total Cholesterol data was available for n=19 “morning”, n= 395 “evening” dosing times.

### Statistics

All statistical analyses (GraphPad Prism Software version 5.03 San Diego, CA) defined significance as α < 0.05. Two group analyses were performed via a two-tailed *t*-test. Analyses of three or more conditions were performed using one-way ANOVA or two-way ANOVAs as indicated. ANOVA analyses were followed by Fisher’s LSD post-hoc to compare between-group differences. Error bars indicate standard error of the mean (SEM).

## Data Availability

All data produced in the present work are contained in the manuscript

## Study approval

All animal procedures were approved by the University of California San Diego IACUC. The human research was reviewed and approved by the VASDHS IRB

## Author Contributions

Conceptualization: OO, RCZ, MJM

Methodology: RCZ, OO, MJM

Investigation: AS, DY, BC, AL, MJM

Writing, Reviewing and Editing: OO, RCZ, MJM

## ACKNOWLEGEMENTS

National Institutes of Health grant R01DK117872 (OO)

Department of Veterans Affairs. BX003431 (MJM)

Larry L. Hillblom Foundation Postdoctoral Fellowship 2019-D-007-FEL (RZ)

## Supplementary Materials

**Supplemental figure 1.**
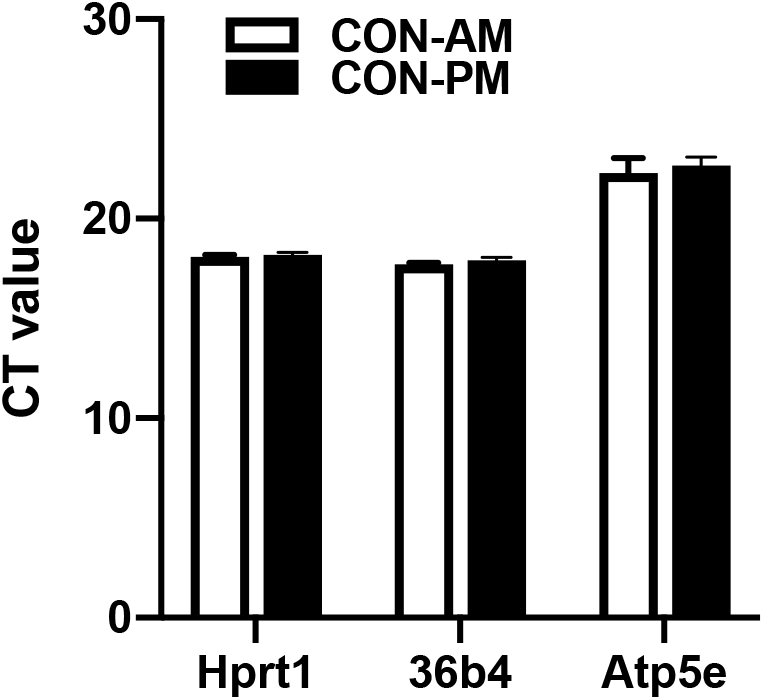
Relative expression of housekeeping genes used for qPCR analysis. **Expression of** Hypoxanthine phosphoribosyltransferase 1 (Hprt1) for the hypothalamus, Acidic ribosomal phosphoprotein P0 (36B4) for the liver and ATP synthase F1 subunit epsilon (Atp5e) for adipose tissues are stable when comparing levels in tissues dissected at ‘AM’ and ‘PM’ timepoints.

## Notes

The authors have declared that no conflict of interest exists

### Competing Interest Statement

The authors have declared no competing interest.

### Author Declarations

The research was reviewed and approved by the VASDHS IRB

